# Testing astrocyte alterations in chronic cocaine users: a longitudinal study using plasma glial fibrillary acidic protein

**DOI:** 10.64898/2026.03.12.26348300

**Authors:** Linus V. Hunglinger, Lukas Eggenberger, Ann-Kathrin Kexel, Bruno Kluwe-Schiavon, Aleksandra Maceski, Markus R. Baumgartner, Jens Kuhle, Erich Seifritz, Boris B. Quednow, Francesco Bavato

**Author notes:** **Corresponding author:** Francesco Bavato, MD, Department of Adult Psychiatry and Psychotherapy, University Hospital of Psychiatry Zurich, University of Zurich, Lenggstrasse 31, CH-8032 Zurich, Switzerland. Equal contribution (Boris B. Quednow, Francesco Bavato).

## Abstract

Preclinical evidence indicates that cocaine exerts acute and chronic effects on astrocyte functioning, which in turn modulate cocaine-related impacts on neural integrity and brain function. However, human evidence for astrocytic involvement in cocaine users (CU) remains limited. Glial fibrillary acidic protein (GFAP) is a marker of astrocyte activation with promising clinical utility in neurological conditions, yet its relevance in the addiction field is unclear. Hence, we investigated plasma GFAP levels in chronic CU (n=41) and cocaine-naive controls (HC; n=34) at baseline and after a 4-month follow-up. GFAP was assessed alongside plasma neurofilament light chain (NfL) levels, a marker of neuroaxonal injury previously associated with cocaine use in the same sample. Contrary to our hypothesis, we found no group differences in plasma GFAP concentrations between CU and HC. Neither cross-sectional nor longitudinal associations between GFAP levels and objective indices of cocaine use (derived from hair testing) were detected. However, exploratory analyses revealed higher plasma GFAP levels among CU with recent cocaine consumption (within the last 7 days), suggesting transient astrocytic responses following acute exposure. Additionally, GFAP and NfL were positively correlated across participants, supporting their functional association. Overall, these findings suggest that while GFAP might not be chronically elevated in CU, it may exhibit transient increases related to recent cocaine use. Further research is warranted to characterize the temporal dynamics and biological significance of these glial responses.

## Introduction

Preclinical evidence indicates that astrocytes might mediate both acute and chronic effects of cocaine on the brain [1]. These glial cells play a critical role in maintaining neuronal homeostasis and in regulating neurovascular coupling, thus modulating both functional activation and long-term survival of neurons [2]. Upon acute exposure to cocaine in mice experiments, astrocytes show functional activation characterised by upregulation of glial fibrillary acidic protein (GFAP) and morphological changes such as hypertrophy and proliferation [3]. Astrocyte activation was recently found to mediate the acute effects of cocaine on cerebral blood flow and neuronal activity in the prefrontal cortex of mice, with chemogenetic astrocytes inhibition preventing these effects [1]. Similarly, changes in synaptic transmission observed in a cocaine self-administration model in rats were attributed to neurons exhibiting increased responsiveness to glutamate released from local astrocytes [4]. Cocaine has also been shown to induce astrocytic autophagy, which correlated with increased GFAP expression and upregulation of proinflammatory factors [5]. This could in turn induce increase oxidative stress and impaired neurotransmitter balance in neurons. Cocaine can also trigger the release of cytoskeletal components such as neurofilament light chain (NfL) [6], which may further activate neuroinflammatory reactions [7]. After repeated cocaine exposure, elevated GFAP immunodensities, which are implicated in astrogliosis, neuroinflammation, and disruption of the blood-brain barrier integrity, have been reported in rodent models, as well as in astrocytic cell culture [5,8-10]. However, tools to investigate in-vivo alterations of astrocyte in humans are so far limited, and the translation of this evidence into clinical research is lacking.

Since the recent introduction of highly sensitive new-generation immunoassay methods, it is possible to measure human blood GFAP levels in a reliable way [11]. In particular, elevated blood GFAP levels have been associated with clinical severity, disease progression, and cognitive impairments in inflammatory and neurodegenerative conditions, such as multiple sclerosis and Alzheimer’s disease [12-14]. On the contrary, data on blood GFAP levels in association with substance use is limited, with preliminary data showing no alterations of GFAP levels in individuals with alcohol use disorder [15].

Therefore, we assessed plasma GFAP levels in chronic cocaine users (CU) and cocaine-naïve healthy control individuals (HC) at baseline and at a 4-month follow-up. GFAP levels were compared with NfL, a well-established marker of active neuroaxonal pathology [16,17] and substance-related brain toxicity [18-20], that has previously shown longitudinal dose– response relationships with cocaine use in this sample [6]. NfL has been shown to correlate with GFAP in neurological samples despite reflecting distinct neurobiological mechanisms [21], supporting the notion that astrocytic activation and neuroaxonal injury often co-occur. We hypothesized that plasma GFAP is elevated in CU at both baseline and follow-up, and that longitudinal changes in GFAP concentrations would be associated with objectively measured changes in cocaine use intensity (employing toxicological hair testing) over this period in a dose-dependent manner. We further hypothesized that plasma GFAP and NfL levels would be interrelated, given the functional connection between astrocyte activation and neuronal integrity [1,4]. Overall, this study aims to advance our understanding of the neurobiological effects of chronic cocaine use on glial components and to clarify the utility of GFAP as a marker of brain pathology in CU.

## Methods

The data were collected at the University Hospital of Psychiatry Zurich as part of the Social Stress Cocaine Study (SSCP) [22,23]. The study was approved by the Research Ethics Committee of the Canton of Zurich (BASEC ID 2016-00278 and 2021-01853). Clinical and laboratory investigations were conducted in strict accordance with the principles of the Declaration of Helsinki. An overlapping dataset was previously published for plasma NfL levels alone [6]. Compared to the previous investigation on NfL (including n=35 CU and n=35 HC previously), the present study included n=41 CU and n=34 HC at baseline. The discrepancy in sample numbers is attributable to differences in blood sample availability, as the current analysis also incorporated all samples for which no follow-up data was available. Of note, NfL levels have been measured again for the present study.

### Participants

General exclusion criteria were: a family history of psychiatric disorders with high genetic risk (h^2^>0.5, e.g., autism, schizophrenia, or bipolar disorder); any severe neurological disorder or brain injury; a current diagnosis of infectious disease or severe somatic disorder; a history of autoimmune, endocrine, or rheumatoid arthritis; intake of medication with potential action on the central nervous system or the physiological stress system during the previous 72h; participation in a large previous study conducted by our lab, the Zurich Cocaine Cognition Study [24,25] and (for women) being pregnant or breastfeeding. Specific inclusion criteria for the CU group included: cocaine as the primary substance of use; a lifetime cumulative consumption of at least 100 g of cocaine, estimated by self-report; and a current abstinence duration of <6 months. Exclusion criteria for CU were: regular use of illegal substances other than cocaine, such as heroin or other opioids (with the exception of cannabis use); polysubstance use pattern according to DSM-IV-TR; and the diagnosis of an axis I adult psychiatric disorder other than cocaine, cannabis, or alcohol abuse or dependence, previous depressive episodes, and attention deficit hyperactivity disorder (ADHD) according to DSM-IV-TR. The exclusion criteria for HC were a DSM-IV-TR axis I adult psychiatric disorder or recurrent illegal substance use (>15 occasions in the lifetime, with the exception of irregular cannabis use). The same inclusion and exclusion criteria have been applied in previous investigations from this study [6].

### Clinical assessment

The clinical interview was carried out by trained psychologists using the Structured Clinical Interview I (SCID-I) in accordance with DSM-IV-TR. Depressive symptoms were assessed using the Beck Depression Inventory (BDI). Symptoms of ADHD were assessed using the ADHD self-rating scale (ADHD-SR).

### Assessment of substance use

For urine analysis, a semi-quantitative enzyme multiplied immunoassay method was used to perform targeting the following substances: amphetamines, barbiturates, benzodiazepines, cocaine, methadone, morphine-related opiates, and delta-9-tetrahydrocannabinol. Liquid chromatography tandem mass spectrometry (LC-MS/MS) was performed to measure substance use in hair over the previous 4 months, as represented in the proximal 4 cm-segment of the hair samples taken from the occiput. In hair, a total of 88 psychoactive substances and substance metabolites were assessed [26]. Cocaine_total_ hair concentration (= cocaine + benzoylecgonine + norcocaine hair concentration in pg/mg hair) was calculated. Cocaine_total_ offers a robust measure discriminating between cocaine incorporation and contamination in hair [27] and can estimate actual cocaine use intensity and risk of dependence [28]. Self-reported substance use was assessed with the structured and standardized Interview for Psychotropic Drug Consumption (IPDC) [29].

### Blood markers

Plasma samples were collected in EDTA tubes and directly centrifuged at room temperature for 15 min at 1500 g. All samples were frozen and stored at −80°C. Plasma GFAP and NfL levels were measured using the Neurology 2-PLEX B array on a single-molecule array (SIMOA) platform. The mean coefficient of variation between intra-assay duplicates was 5.15% for NfL and 9.24% for GFAP.

### Statistical analysis

All statistical analyses were computed with R (Version 4.5.0). Quantitative variables were tested for normal distribution using the Shapiro-Wilk test and logarithmic transformed if necessary. Between-group comparisons (CU/HC) of sociodemographic and clinical data were performed using Student’s t-tests (for normally distributed quantitative data), Mann– Whitney U-tests (for non-normally distributed quantitative data), or Pearson’s chi-square test (for categorical data). Linear mixed-effect models (LMMs) were used to estimate the effects of group (CU vs. HC), timepoint (baseline vs. 4-month follow-up), and their interaction on GFAP and NfL levels, with age, sex, and BMI as covariates and subject as a random effect. Age was included due to the strong association between advancing age and increased NfL and GFAP levels while BMI was included due to the reported negative association with NfL and GFAP levels [30]. Sex was included to correct for potential sex-specific effects of cocaine use on brain pathology [31], and because of the reported sex-effect on GFAP levels [11]. An LMM was also used to assess GFAP effects on NfL levels, with timepoint, group, age, sex and BMI as covariates and subject as a random effect. Spearman’s correlation coefficients were calculated to investigate dose–response relationships between substance use variables and blood markers, both cross-sectionally (baseline and follow-up levels; *n*=41 and *n*=30) and longitudinally (changes from baseline to follow-up; *n*=29) in CU. For testing single group differences, Tukey’s Honestly Significant Difference (HSD) Post-hoc Tests were applied, controling the family-wise error rate (Type I error). The significance level was set at *p*<.05 (two-tailed).

## Results

### Sociodemographic and clinical characteristics

Sociodemographic and clinical data is reported in Table 1. CU were slightly older than HC, even though no significant group difference occurred. The group comparisons for blood markers were nevertheless corrected for age. Higher self-rating scores for ADHD and depression, higher cannabis and alcohol use, and lower education in CU are consistent with previous publications from this sample [22,23], and with similar reports in other studies [32]. Detailed substance use patterns are summarized in the supplementary materials (Table S1).

**Table 1.** Sociodemographic and clinical variables.

| Baseline |  |  |  |  |  |
| --- | --- | --- | --- | --- | --- |
|  | Overall<br>(*n*=75) | Control<br>(*n*=34) | Cocaine<br>(*n*=41) | *p*-value | SMD |
| **Female sex, *n* (%)** | 28 (37.3) | 12 (35.3) | 16 (39.0) | .926 | 0.077 |
| **Age, mean (SD)** | 31.2 (7.4) | 29.6 (7.2) | 32.5 (7.4) | .090 | 0.399 |
| **BMI, mean (SD)** | 24.4 (3.7) | 23.5 (3.1) | 25.1 (3.9) | .058 | 0.450 |
| **School education, years** | 10.0 (1.3) | 10.5 (1.5) | 9.5 (1.0) | **.001<sup>\*\*\*</sup>** | 0.777 |
| **ADHD_SR, score** | 12.9 (9.7) | 9.9 (7.7) | 15.4 (10.5) | **.013<sup>\*</sup>** | 0.601 |
| **ADHD, DSM (yes/no)** | 16 (21.3) | 5 (14.7) | 11 (26.8) | .321 | 0.302 |
| **History of MDD, DSM (yes/no)** | 17 (22.7) | 3 (8.8) | 14 (34.1) | **.020<sup>\*</sup>** | 0.648 |
| **BDI, score** | 6.2 (6.4) | 4.2 (5.5) | 7.7 (6.8) | **.018<sup>\*</sup>** | 0.566 |
| **Cocaine** |  |  |  |  |  |
| cocaine<sub>total</sub>, pg/mg in hair | — | — | 28,660<br>(63,579) | — | — |
| grams per week | — | — | 1.5 (1.9) | — | — |
| abstinence days | — | — | 23.4 (43.3) | — | — |
| < 7 days, *n* (%) | — | — | 24 (58.5) | — | — |
| ≥7 days, *n* (%) | — | — | 17 (41.5) | — | — |
| **Alcohol, grams per week** | 101.6 (82.8) | 79.3 (75.0) | 120.5 (85.3) | **.032<sup>\*</sup>** | 0.512 |
| **Cannabis, grams per week** | 0.28 (1.53) | 0.08 (0.17) | 0.44 (2.04) | .341 | 0.248 |
| **Nicotine, cigarettes per week** | 73.3 (60.1) | 60.9 (47.9) | 82.8 (67.1) | .128 | 0.376 |
| **Blood markers, mean (SD)** |  |  |  |  |  |
| GFAP, pg/mL | 40.9 (17.8) | 39.2 (15.1) | 42.4 (19.9) | .430 | 0.186 |
| NfL, pg/mL | 5.9 (3.2) | 4.9 (1.8) | 6.8 (3.8) | **.010<sup>\*</sup>** | 0.633 |
| Follow-Up |  |  |  |  |  |
|  | Overall<br>(*n*=58) | Control<br>(*n*=28) | Cocaine<br>(*n*=30) | *p*-value | SMD |
| **BMI, mean (SD)** | 24.1 (3.8) | 23.5 (3.5) | 24.7 (3.9) | .231 | 0.319 |
| **Cocaine** |  |  |  |  |  |
| cocaine<sub>total</sub>, pg/mg in hair | — | — | 20,433<br>(30,637) | — | — |
| grams per week | — | — | 1.6 (2.1) | — | — |
| abstinence days | — | — | 33.8 (69.1) | — | — |
| < 7 days, *n* (%) | — | — | 10 (33.3) | — | — |
| ≥7 days, *n* (%) | — | — | 20 (66.7) | — | — |
| **Blood markers, mean (SD)** |  |  |  |  |  |
| GFAP, pg/ml | 48.7 (16.3) | 48.7 (15.9) | 48.6 (17.0) | .981 | 0.006 |
| NfL, pg/ml | 6.5 (4.3) | 5.3 (1.9) | 7.7 (5.4) | **.033<sup>\*</sup>** | 0.582 |

Note. *n*=number of participants. SMD=standardized mean difference.

### Group comparison of plasma GFAP and NfL levels, and GFAP-NfL association

#### GFAP

No group effect was observed for plasma GFAP levels after correction for timepoint, age, BMI, sex (raw plasma GFAP levels CU vs. HC: mean ± SD: 45.04 ± 18.83 vs. 43.46 ± 16.07 pg/mL; median [IQR]: 40.37 [31.99–55.55] vs. 41.40 [33.32–54.94] pg/mL; LMM group effect: β=0.09, 95% CI [-0.10, 0.27], *t*(90)=0.90, *p*=.370; c.f. Figure 1a). In the same model, no influence of age (β=0.01, 95% CI [-0.01, 0.01], *t*(71)=0.52, *p*=.608), sex (β=0.07, 95% CI [-0.11, 0.26], *t*(70)=0.78, *p*=.440), or BMI (β=-0.02, 95% CI [-0.04, 0.01], *t*(69)=-1.30, *p*=.198), on plasma GFAP levels was observed, while a positive effect of timepoint was found (β=0.23, 95% CI [0.11, 0.35], *t*(57)=3.72, *p* <.001).

**Figure 1.**
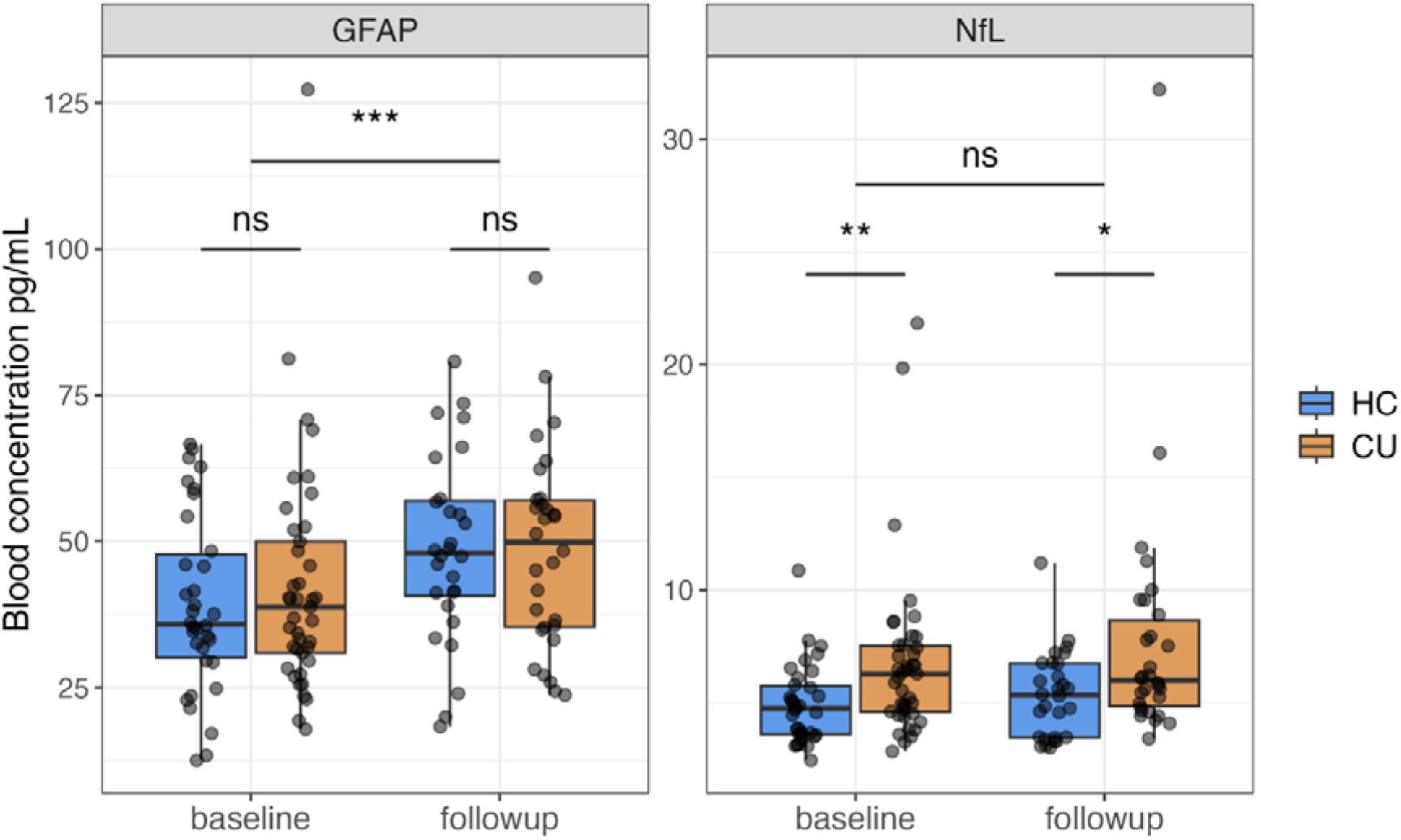
Group comparison of GFAP and NfL levels. Boxplots showing individual plasma GFAP and NfL levels. Central horizontal lines indicate median values; boxes illustrate the ranges between lower and upper quartiles. Abbreviations: HC: Healthy Controls; CU: Cocaine Users; NfL: Neurofilament Light Chain; GFAP: glial fibrillary acidic protein; P-values are reported for Tukey’s HSD post-hoc test. ns=not significant; * p < 0.05, ** p < 0.01, *** p < 0.001.

#### NfL

Plasma NfL levels (which were measured again for this study) were increased in CU compared to HC (raw plasma NfL levels CU vs. HC: mean ± SD: 7.16 ± 4.54 vs. 5.09 ± 1.83 pg/mL; median [IQR]: 6.12 [4.66–7.84] vs. 4.85 [3.50–6.13] pg/mL; LMM group effect: β=0.26, 95% CI [0.09, 0.43], *t*(97)=2.96, *p*=.004; c.f. Figure 1b), consistent with our previous report from an overlapping sample [6]. Age and BMI were found to be significant covariates (age: β=0.02, 95% CI [0.01, 0.03], *t*(65)=3.99, *p* <.001; BMI: β=-0.03, 95% CI [-0.05, -0.01], *t*(62)=-2.60, *p*=.012), while sex and timepoint had no relevant impact in the model (sex: β=0.06, 95% CI [-0.10, 0.23], *t*(64)=0.74, *p*=.461; time: β=0.09, 95% CI [-0.05, 0.22], *t*(54)=1.28, *p*=.205).

Finally, a positive association between plasma NfL and GFAP levels was observed (LMM GFAP effect on NfL levels: β=0.31, 95% CI [0.15, 0.46], *t*(117)=3.75, *p* <.001). Longitudinal changes in GFAP levels (ΔGFAP) were positively associated with longitudinal changes in NfL levels (ΔNfL) (ρ=.28, *p*=.036, *n*=56, see supplementary figure 1).

### Associations between plasma GFAP and substance use variables

Among cocaine users, plasma GFAP levels were not associated with the hair concentration of cocaine_total_ (baseline: ρ=.20, *p*=.203; follow-up: ρ=.12, *p*=.539) neither with self-reported cocaine consumption per week in the last 6 months (baseline: ρ=.28, *p*=.083; follow-up: ρ=.07, *p*=.707). Longitudinal changes in cocaine consumption (Δcocaine_total_) showed no association with ΔGFAP (ρ=.24, *p*=.213).

In an exploratory analysis, the role of abstinence on GFAP levels was tested (see Figure 2). This analysis was motivated by recent evidence suggesting the elimination time of GFAP in blood to be within 150-200 hours [33]. Here, a significant effect of abstinence on GFAP levels was observed after correction for timepoint, age, sex, and BMI (LMM, abstinence effect on GFAP: β=-0.10, 95% CI [-0.17, -0.03], *t*(64)=-2.78, *p*=.007). In particular, CU reporting cocaine use within 7 days had higher GFAP levels compared to HC (mean [median] ± SD, pg/ml: 50.51 [42.20] ± 20.97 vs. 43.46 [41.40] ± 16.07; LMM group effect: β=0.22, 95% CI [0.02, 0.42], *t*(56)=2.11, *p*=.039), while CU with no cocaine use within 7 days, had comparable GFAP levels than HC (mean [median] ± SD, pg/ml: 40.01 [35.30] ± 16.30 vs. 43.46 [41.40] ± 16.07; LMM group effect: β=-0.13, 95% CI [-0.35, 0.08], *t*(79)=-1.15, *p*=.252).

**Figure 2.**
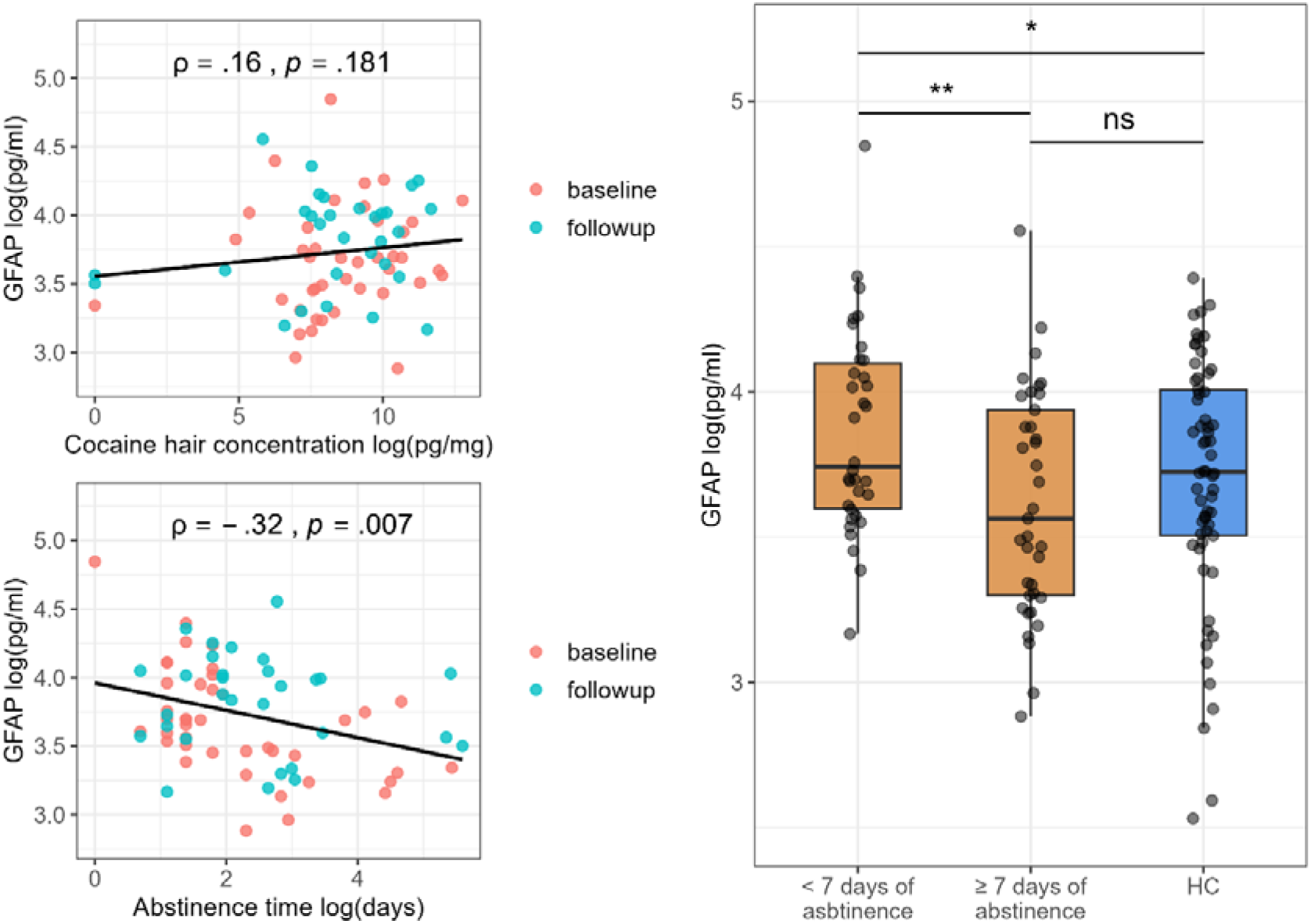
Associations between GFAP and substance use variables. A) Scatterplots showing the relationship between hair concentration of cocaine_total_ and abstinence days with plasma GFAP. B) Boxplots showing individual plasma GFAP (log-transformed) for CU patients with or without cocaine use within the last 7 days. Central horizontal lines indicate median values; boxes illustrate the ranges between lower and upper quartiles. Abbreviations: HC: Healthy Controls; CU: Cocaine Users; GFAP: glial fibrillary acidic protein; P-values are reported for Tukey’s HSD post-hoc tests. ns=not significant; * p < 0.05, ** p < 0.01, *** p < 0.001.

## Discussion

A link between cocaine use and astrocyte alterations has long been suggested in animal models but *in vivo* evidence in humans was lacking. In this study, we show that plasma levels of the astrocyte marker GFAP do not differ between recently abstinent CU and HC overall. However, an exploratory observation about increased GFAP levels within the last 7 days of cocaine consumption, suggesting a transient alteration related to acute effects. We also provide evidence of an association between GFAP and the neuraxonal integrity marker NfL in CU.

An increase of blood GFAP levels have been reported in numerous neurological conditions such as traumatic brain injury, multiple sclerosis, and Alzheimer’s disease [11]. Here, GFAP elevation has been shown to reflect the degree of clinical severity and structural brain pathology and to hold prognostic utility for clinical outcomes [13,34]. Notably, after mild traumatic brain injuries, GFAP levels have been found to reach peak concentration within 24 hours normalize after 7 days [33]. In psychiatry, elevated GFAP levels have been reported in the cerebrospinal fluid and blood of patients with major depressive disorder, bipolar disorder, and treatment-resistant schizophrenia, while no alteration was observed in patients with alcohol use disorder [15,35-39].

In the case of cocaine, evidence of associations between chronic use and GFAP alterations are mostly limited to preclinical studies. Fattore et al. showed increase in GFAP immunostaining in the mouse dentate gyrus directly after acute and chronic cocaine treatment [8]. Scofield and colleagues reported reduced GFAP levels in the nucleus accumbens of rats following extinction of cocaine self-administration [40]. A concurrent reduction in the number of GFAP labelled astrocytes was also observed. Accordingly, a biphasic effect of cocaine on astrocytes with acute GFAP overexpression followed by post-acute functional deactivation (or reduced clearance) leading to decreased GFAP release in blood can be hypothesized. This is also consistent with magneto-spectroscopy evidence of dynamic alterations of the striatal glutamate homeostasis in CU, which is crucially regulated by astrocyte functioning [41-43]. On the contrary, a postmortem study in CU after cocaine overdose reported no effects on brain GFAP immunopositivity, although this investigation entailed limited semiquantitative analysis and suboptimal topological characterization [44]. Thus, our findings provide novel evidence on *in vivo* GFAP involvement in CU and are overall consistent with preclinical evidence of time-dependent astrocyte alterations following cocaine use. Finally, our data also parallels observations in mild traumatic brain injury, where blood GFAP levels normalize within 7 days [33].

The biological meaning of blood GFAP levels in neurological and psychiatric conditions is still debated. While it was initially suggested that GFAP release into the blood stream occurs as a consequence of astrocyte injury or death, current evidence hint at different possible mechanisms involved [11]. Functional activation, reactive astrogliosis, or structural reorganization might also mediate a peripheral increase of GFAP levels [45]. On the contrary, GFAP reduction in blood might reflect reduced astrocyte activity [40,46] or rebound mechanisms following acute GFAP release. Intriguingly, the association between GFAP and NfL levels in the CU sample might suggest a link between astrocyte changes and NfL elevation, which is consistent with preclinical evidence of astrocyte mediation in cocaine-induced neural pathology [1]. Conversely, NfL release has recently been linked to the activation of inflammatory pathways in microglia, which could in turn lead to secondary astrocyte activation and increased GFAP expression [7]. Clearly, these mechanistic speculations still lack of robust experimental validation and would require further observations with dense sampling to fully capture its temporal dynamics.

From a clinical perspective, the lack of overall group effects for GFAP might limit its validity for diagnostic purposes and cross-sectional patient stratification. The limited relevance of GFAP for diagnostic uses on an individual level is not surprising considering the limited group-level effect sizes even in clinically stable neurological conditions [11], similarly to NfL [47]. Nonetheless, the large effect size of state-dependent effects might still open opportunities for neurobiological investigations on in-vivo astrocyte involvement in CU and might have relevant implications in the longitudinal monitoring of cocaine-induced brain effects. Moreover, our findings warrant careful account of recent substance use in future studies addressing GFAP in different medical conditions.

This study bears some relevant limitations. 1) The sample size is small, particularly in relationship to abstinence subgroups. Future research should focus on replicating these findings in larger and more diverse populations to enhance generalizability and include more participants with recent cocaine use. Neuroimaging investigations should also confirm if GFAP alterations reflect structural brain alterations as observed in neurological conditions. 2) The severity of cocaine use was broadly distributed in our chronic CU, spanning from non-dependent regular use to very intense consumption patterns. While such a large variance in cocaine use severity is beneficial for the detection of dose-response correlations, it may has limited the detection of overall group differences. 3) We observed a small but statistically significant time effect for GFAP, whereas no such effect was detected for NfL. While the reason for this discrepancy remains unclear, a subtle batch effect on GFAP measurements cannot be fully excluded despite not being apparent for NfL. Importantly, all statistical analyses were adjusted for timepoint, reducing the likelihood that this influenced the main findings.

In summary, our findings do not support the notion of sustained astrocytic activation in chronic CU, as indexed by plasma GFAP levels, which did not differ from HC either cross-sectionally or over a 4-month follow-up. Moreover, GFAP concentrations were not associated with objective measures of cumulative cocaine exposure, suggesting that cocaine use intensity is not accompanied by persistent systemic astrocytic alterations detectable in peripheral blood. However, exploratory analyses indicating elevated GFAP levels among users with recent cocaine intake point to a potential transient astrocytic response to acute cocaine exposure. The observed positive association between GFAP and NfL across participants further aligns with their coupled roles in neuroglial and neuroaxonal processes. Together, these results suggest that astrocytic involvement in cocaine use may be dynamic rather than chronically elevated, underscoring the need for future studies with finer temporal resolution to clarify the short-term glial responses to cocaine and their implications for brain health in people with cocaine use disorder.

## Data Availability

All data produced in the present study are available upon reasonable request to the authors

## Acknowledgements

This study was supported by a grant from the Swiss National Science Foundation (Grant Nr. 105319_162639) to BBQ. BKS received a grant from the Coordination for the Improvement of Higher Education Personnel, CAPES, Brazil (Grant Nr.: 99999.001968/2015-07). FB received salary funding from the Swiss National Science Foundation (Grant Nr: 217637). The funders had no role in the study design, in the collection, management, analysis, and interpretation of data, in the preparation, review, and approval of the manuscript, or in the decision to submit the manuscript for publication. We are grateful to Monika Näf, Chantal Kunz, Marlon Nüscheler, Selina Maisch, Jocelyn Waser, Anna Burkert, Meret Speich, Maxine de Ven, Zoé Dolder, Zoe Hillmann, Jessica Grub, and Priska Cavegn for their excellent support with recruitment and assessment of the participants.

## Conflict of interest

The authors declare no conflict of interest.

## Author contributions

BBQ and FB developed the study concept and design. BBQ and FB rose the funding for the study and the present analysis. ES contributed additional funding. AKK and BKS conducted assessments. AM and JK provided GFAP and NfL plasma analyses and supported data interpretation. MRB conducted hair analyses and supported data interpretation. LVH and LE conducted the statistical analyses. LVH drafted the first manuscript. All authors revised and approved the final manuscript. BBQ and FB provided supervision.

## Notes

### Competing Interest Statement

The authors have declared no competing interest.

### Author Declarations

The study was approved by the Research Ethics Committee of the Canton of Zurich (BASEC ID 2016-00278 and 2021-01853). Clinical and laboratory investigations were conducted in strict accordance with the principles of the Declaration of Helsinki.

